# The Reliability and Validity of the Hebrew Patient Health Questionnaire (PHQ-9) in the General Population

**DOI:** 10.1101/2021.07.13.21260485

**Authors:** Tomer Yona, Asaf Weisman, Uri Gottlieb, Eshed Lin, Youssef Masharawi

**Author notes:** **Corresponding author** Tomer Yona, Technion, Israel Institute of Technology, Haifa, Israel.

## Abstract

**Objective:** To assess the psychometric properties of the Hebrew version of the Patient Health Questionnaire (PHQ-9) in the general population.

**Methods:** Using an online survey, we assessed test-retest reliability with a two-week time interval. A total of 118 participants enrolled in the study, of whom 103 completed the survey twice. Each participant filled out the PHQ-9 and the 12-Item Short Form Survey (SF-12). Our statistical analysis includes Cronbach’s alpha, Intraclass Correlation Coefficient (ICC2,1), Spearman’s rank correlation coefficient, Standard Error of Measurement (SEM), and Minimal Detectable Change (MDC).

**Results:** Internal consistency of the Hebrew version of the PHQ-9 ranged from α=0.79-0.83. The test-retest reliability of the questionnaire is good (ICC2,1= 0.81), and it is moderately and negatively correlated to the mental component of the SF-12 (Spearman ρ= -0.57, p< .05). The SEM of the PHQ-9 is 1.83 points, and the MDC was found to be 5 points.

**Conclusion:** The Hebrew version of the PHQ-9 is valid and reliable for screening self-reported depressive symptoms online in the general Hebrew-speaking population.

## INTRODUCTION

Depression is among the leading causes of disability worldwide. Additionally, the prevalence of depression and depressive symptoms has been rising in recent decades.^1,2^ Despite the availability of many self-reported tools to screen and diagnose perceived depressive symptoms, depression remains under-detected among various populations.^3-5^

The Patient Health Questionnaire (PHQ-9) was developed as a short questionnaire based on the Diagnostic and Statistical Manual of Mental Disorders (DSM-IV) criteria to diagnose depressive disorders.^6^ It aims to screen self-reported depressive symptoms is freely available and has been translated and validated in many different languages and populations.^7-10^ However, the psychometric properties of the Hebrew version of the PHQ-9 have not been investigated.

Properly validated and reliable tests are essential for clinical decisions, diagnoses, and prognoses.^11^ Moreover, using unvalidated or unreliable instruments can affect how authors report on a study’s results.^12^ Consequently, this paper aims to assess the internal consistency, test-retest reliability, and construct validity of an online administration of the Hebrew version of the PHQ-9 on the general Hebrew-speaking population in Israel.

## METHODS

Tel Aviv University’s ethics committee approved this study (Number 0003314-2). The study design followed the COSMIN study design checklist for patient-reported outcome measurement instruments.^13^

### Procedures

We used an online platform (www.Alchemer.com) to disseminate the survey in June 2021. Hebrew-speaking respondents were recruited from the Israeli adult general population using social media. The first page of the survey consisted of the survey description, the informed consent, and the researcher’s contact details. Inclusion criteria were a minimum age of 18 years old and the ability to read and comprehend Hebrew at a native level of language proficiency.

Each respondent completed the survey twice, with a two-week interval between testing. The first time-point included only the PHQ-9 and the second time-point included the 12-Item Short Form Survey (SF-12) and the PHQ-9. Additionally, at the second time point, respondents were first asked whether they “experienced a change in their health in the previous two weeks?” Any respondent that answered “yes” was directed out of the survey and was excluded from the reliability analysis.

### Patient Health Questionnaire (PHQ-9)

The PHQ-9 is a self-reported questionnaire consisting of nine items to assess the severity of depressive symptoms. It is used to determine the presence of bothersome symptoms experienced by the participant in the last two weeks using a 0-3 Likert scale. The minimum total score (0) indicates no depressive symptoms, and the highest total score (27) indicates severe depressive symptoms. We used the official Hebrew version of the PHQ-9, freely available online (https://www.phqscreeners.com).

### The 12-Item Short Form Survey (SF-12)

The SF-12 is a self-reported questionnaire consisting of twelve items to assess the respondent’s general, physical, and mental health. The summary score of the SF-12 consists of a Physical Composite Scale (PCS) and a Mental Health Composite Scale (MCS). A higher score indicates a better health status. To assess the validity of the PHQ-9, we used the Hebrew version of the SF-12 that has already been proven as reliable and valid.^14^

### Statistical analysis

Statistical analyses were performed using IBM SPSS Statistics for Windows (Version 25, Armonk, NY) and Jamovi (Version 1.6, The Jamovi project). Internal consistency was calculated using Cronbach’s alpha (α), while Intraclass Correlation Coefficient (ICC2, 1) was used to calculate the test-retest reliability and Spearman’s rank correlation coefficient (ρ) was used for construct validity. The following scale was used to determine ICC: higher than 0.90 - excellent reliability; above 0.75 - good reliability; and 0.50-0.75 - moderate reliability.^15^ For Spearman’s correlations, the following scale was used: 0.7-0.9= strong; 0.4-0.6= moderate; and 0.1-0.3= weak.^16^ For the calculation of the Standard Error of Measurement (SEM), the pooled standard deviations formula was used 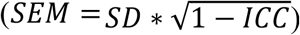. The formula for the Minimal Detectable Change (MDC) was 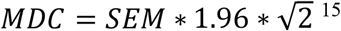. Lastly, we considered a floor and ceiling effect if more than 15% of participants achieved the highest or lowest score in the PHQ-9.^17^

## RESULTS

A total of 132 respondents entered the survey in the first time point, of whom 118 respondents completed the survey. In the second time point, 113 participants completed the survey, of whom 6 respondents were excluded because they stated that their health status changed in the last two weeks, and 4 respondents were lost to follow-up, leaving 103 respondents in the reliability analysis.

The age of respondents ranged from 24 to 77 years of age, with 50.0% self-identifying as female and 50.0% as male. The majority reported having completed higher education (106 participants, 89.8%; see Table 1). The mean score of the PHQ-9 was 4 points, the minimum score was 0, and the maximum score was 19 points.

**Table 1.** Characteristics of the participants.

|  | **n=118** |
| --- | --- |
| **Age (years)** | 40.2 ± 10.8 |
| **Gender (%)** |  |
| Male | 59 (50.0) |
| Female | 59 (50.0) |
| **Marital status (%)** |  |
| Single | 14 (11.9) |
| In a relationship | 25 (21.2) |
| Married | 78 (66.1) |
| Divorced | 1 (0.8) |
| **Education level (%)** |  |
| High school/Technical school | 10 (8.6) |
| Higher education | 106 (89.8) |
| Missing | 2 (1.7) |

The internal consistency of the PHQ-9 was α=0.79 for the first time-point and 0.83 for the second time-point. We identified a good test-retest reliability (ICC2, 1= 0.81), a SEM score of 1.83 points, and a MDC score of 5.07 points. The PHQ-9 moderately and negatively correlated to the SF-12’s MCS (Spearman ρ= -0.57, p< .05), but not to the PCS (Spearman ρ= -0.12, p= .22; see Table 2). None of the respondents scored the highest score in the PHQ-9, while seven respondents (6.8%) scored zero.

**Table 2.** Psychometric properties of the PHQ-9 questionnaire.

|  | **n=103** |
| --- | --- |
| **Cronbach's $\alpha$** | 0.79-0.83 |
| **ICC** | 0.81 |
| **SEM** | 1.83 |
| **MDC** | 5.07 |

ICC = Intraclass Correlation Coefficient. SEM = Standard Error of Measurement. MDC = Minimal Detectable Change.

## DISCUSSION

This study investigated the psychometric properties of the Hebrew version of the PHQ-9. We found that the questionnaire is reliable and valid for use among the Hebrew speaking adult population.

The internal consistency of the Hebrew version of the PHQ-9 (α=0.79-0.83) was slightly lower than the one reported by the developers of the original, English version of the PHQ-9 (α=0.86-0.89)^6^ and on par with other translated language versions (α=0.78-0.86).^10,9,7^ Internal consistency describes if the items on a questionnaire measure the same general construct, and a Cronbach’s alpha of 0.8 is considered acceptable.^15^

We identified a good test-retest reliability (ICC= 0.81), while other versions reported a reliability of 0.59-0.87. The wide range of reliability may be due to different populations (general population, HIV, breast cancer), different languages (Chinese, Portuguese, Swahili, English), different time intervals (one-eight weeks), and different sample sizes (45-187 participants).^9,8,7^ We chose a two-week time interval and 103 participants as recommended by the COSMIN guidelines.^11,13^

The construct validity of the PHQ-9 is varied as different authors chose different comparable questionnaires. We chose the SF-12, as it was previously validated in Hebrew.^14^ We found the SF-12 MCS is moderately and negatively correlated with the PHQ-9 (Spearman ρ= -0.57, p< .05). Others used various SF questionnaires and reported on varying correlations from -0.43 to -0.73^7,18,19,6^ between the PHQ-9 and the mental aspect of the SF questionnaires. The differences between the studies may be due to different clinical and non-clinical populations.

Another key point is that the respondents in our study completed the PHQ-9 online. Yet, the psychometric properties of the questionnaires are on par with previous studies which administrated the survey by mail, phone, or in-person as a paper questionnaire.^18,19,7,20^ Consequently, we conclude the PHQ-9 can be administered online.

Our study is not without limitations. Firstly, most participants had a higher education degree, limiting the generalizability of our results to the general population. Secondly, we did not compare the PHQ-9 to a psychiatric evaluation. Thirdly, the mean PHQ-9 score of the participants was 4 points, and the highest was 19 points. Hence, people with increased depressive symptoms were underrepresented. Further studies could assess the Hebrew version of the PHQ-9 in a more varied population and compare it to a psychiatric evaluation to determine both sensitivity and specificity.

## CONCLUSION

The Hebrew version of the PHQ-9 is valid and reliable for screening self-reported depressive symptoms among the Hebrew-speaking general population. Further, it is feasible to administer it online.

## Data Availability

Data available on request from the authors

